# High-Sensitivity Near Point-of-Care Detection of Asymptomatic and Sub-microscopic Plasmodium Infections in African Endemic Countries

**DOI:** 10.1101/2025.07.12.25331425

**Authors:** Dimbintsoa Rakotomalala Robinson, Ivana Pennisi, Matthew L. Cavuto, Francois Kiemde, Martin Chamai, Diane Yrgnur Some, Elliot Quigley, Kenny Malpartida-Cardenas, Mamadou Ousmane Ndiath, Simon Correa, Bubacarr Darboe, Lindsay B Stewart, Pantelis Georgiou, Mamadu Baldeh, Halidou Tinto, Aubrey J. Cunnington, Annette Erhart, Umberto D’Alessandro, Jesus Rodriguez-Manzano, NIHR Global Health Research Group on Digital Diagnostics for African Health Systems

**Author notes:** These authors contributed equally (Dimbintsoa Rakotomalala Robinson, Ivana Pennisi, Matthew L. Cavuto).

## Abstract

The limited diagnostic capacity to detect asymptomatic individuals with low parasite densities continues to hinder malaria elimination efforts across Africa. We adapted a near point-of-care LAMP-based diagnostic platform, originally designed for viral detection in respiratory and skin lesion specimens, for malaria diagnosis using capillary blood. The resulting Pan*/Pf* malaria test meets the Malaria Eradication Research Agenda (malERA) essential criteria for community-level malaria screening, with an analytical limit-of-detection of 0.6 parasites/μL and a sample-to-result turnaround time under 45 minutes. We evaluated the test using 672 capillary blood samples obtained via finger pricks from individuals enrolled at the community level in The Gambia and Burkina Faso, including 146 positives for *P. falciparum* confirmed by dried blood spot qPCR. The test achieved a sensitivity of 95.2% [95% CI: 90.4-98.1] and specificity of 96.8% [95% CI: 94.9-98.0]. It also detected 94.9% (130/137) of asymptomatic malaria infections and 95.3% (41/43) of sub-microscopic cases (<16 parasites/µL), outperforming expert light microscopy, which detected 70.1% (96/137) and 0% (0/43), and rapid diagnostic tests, which detected 49.6% (68/137) and 4.7% (2/43), respectively. This field molecular method represents a sensitive and scalable diagnostic solution with the potential to support test-and-treat strategies for malaria elimination across Africa.

## INTRODUCTION

Despite substantial investment, malaria remains a significant global health concern, with an estimated 263 million cases and 597,000 deaths reported in 2023.^1^ The African continent bears the highest burden, accounting for 94% of malaria cases and 95% of deaths, predominately caused by *Plasmodium falciparum*.^1^ While high transmission settings rely on universal coverage of standard interventions such as insecticide-treated nets (ITNs), indoor residual spraying (IRS), intermittent preventive treatment for pregnant women (IPTp), and seasonal malaria chemoprevention (SMC), regions with declining transmission urgently require innovative strategies to accelerate malaria elimination.^2–4^

To accelerate progress, it is crucial to target the entire human reservoir of infection, including both symptomatic and asymptomatic *Plasmodium*-infected individuals.^5^ Passive case detection at health facilities identifies clinical malaria cases while asymptomatic carriers in the community are unlikely to seek medical care and remain undetected. In contrast, active detection interventions (ADIs), involving community-level test-and-treat strategies, offer a promising approach to addressing this gap.^5,6^ ADIs also have the advantage of avoiding the widespread drug exposure in Mass Drug Administration (MDA), a strategy that treats entire populations regardless of the infection status. MDA can lead to several challenges, including the risk of drug resistance, the unnecessary treatment of uninfected individuals, and the logistical burden of implementing large-scale treatments.^7,8^ However, the success of ADIs hinges on the availability of highly sensitive diagnostic tools capable of detecting all or most malaria infections.^2,8,9^

Asymptomatic carriage is often associated with low parasite densities, typically below 100 parasite/µL, and frequently fall under the detection threshold of conventional diagnostic methods such as light microscopy (LM) and rapid diagnostic tests (RDTs).^5,10,11^ These infections, termed sub-microscopic or sub-patent, occur across the whole spectrum of malaria transmission intensity with the highest proportion (60-70%) among infected individuals in low-transmission settings.^12^ Despite their low density, sub-microscopic infections contribute to ongoing transmission by harbouring gametocytes that can infect mosquitoes.^13,14^ RDTs and LM have a limit of detection (LOD) around 100-200 and 50-100 parasites/µl, respectively.^15^ Both tests face challenges in reliably identifying asymptomatic carriers, particularly those with low parasitaemia. LM could be subjective depending on the skills of the microscopist, whereas the increased number of reported deletions of *PfHRP2* and *PfHRP3* genes are compromising the use of HRP-based RDTs.^1^ In contrast, Polymerase Chain Reaction (PCR) and quantitative PCR (qPCR) detect infections at densities as low as 0.002 parasites/µl, but their widespread adoption faces significant challenges due to the complexity of execution, particularly in resource-constrained settings.^16,17^ PCR-based techniques requires labour-intensive procedures, substantial costs, advanced laboratory infrastructure, and highly skilled technicians. Additionally, results can take several hours or even days to produce, significantly delaying diagnosis and treatment.^17^ Therefore, there is a pressing need for a diagnostic technology that combines both the high analytical sensitivity of molecular methods with the practicality and accessibility required for widespread use in resource-constrained field settings currently achieved by RDTs. As alternatives to standard PCR, several Nucleic Acid Amplification Tests (NAATs) have emerged, including Nucleic Acid Sequence-Based Amplification (NASBA), Recombinase Polymerase Amplification (RPA) and Loop-mediated isothermal amplification (LAMP).^18–21^

LAMP-based technologies offer a promising alternative, combining the high sensitivity of molecular diagnostics with simpler equipment and operational requirements.^19,22^ Unlike PCR, LAMP allows the amplification of target nucleic acid sequences at a constant temperature, an advantageous feature for field deployment, enabling the use of less expensive and more portable battery-powered block heaters. Unfortunately, similar to PCR, to achieve sufficient sensitivity LAMP requires high quality nucleic acid extraction to be performed prior to amplification.^19^ As a result, many LAMP assays still rely on lengthy, multi-step nucleic acid extraction kits, making them less practical for community-based screening that demands high throughput and a shorter time-to-result.^23–25^ Moreover, liquid LAMP reagents, as for other molecular approaches, require cold-chain storage which represents a challenge for deployment in resource-constrained settings. To our knowledge, only two LAMP-based malaria diagnostic platforms are commercially available, the Loopamp^TM^ Malaria Detection Kits (Eiken Chemical Co., Tokyo, Japan)^26^ and the Alethia® Malaria (Meridian Bioscience Inc., Cincinnati, OH, USA), previously called Illumigene®^27^. While both platforms eliminate the need for a cold chain through lyophilisation of the LAMP reagents, they also employ shortened sample preparation processes which may increase reaction inhibition and lower nucleic acid recovery. Both of them also rely on instruments that measure turbidimetry or fluorescence emission for result readout, increasing the cost and bulk of each solution.^25^ In recent years, the Alethia® Malaria assay has been widely deployed in non-endemic high-income countries for the diagnosis of malaria in returning travellers, due to its high diagnostic accuracy relative to RDTs.^28–30^ However, while the Alethia® platform maintains relative ease-of-use when compared to standard laboratory-based molecular methods, the cost of the instrument (> $20,000) and limited throughput (< 10 samples per instrument every 40 minutes) remain significant barriers to its wider adoption and remote deployment in rural African settings.

To address the challenges outlined above, we present a novel near point-of-care (POC) molecular approach for detecting *Plasmodium* genes. This solution, combining the magnetic bead-based nucleic acid extraction technology from SmartLid and the lyophilised colourimetric LAMP chemistry from the Dragonfly platform (originally developed for detecting viral respiratory and skin infections)^31,32^, was optimised for medium to high-throughput malaria testing using capillary blood samples obtained via finger pricks in resource-limited settings. First, the analytical performance of this method was compared against the Alethia® Malaria Test, dried blood spot (DBS)-qPCR, and whole blood qPCR (WB-qPCR) using *Plasmodium* culture spiked into whole blood. Next, its clinical performance was evaluated in standard laboratories at the Medical Research Council (MRC) Unit The Gambia at LSHTM and at The Clinical Research Unit of Nanoro. This was achieved by collecting capillary blood samples from individuals enrolled in a community-based survey in rural Burkina Faso and The Gambia, benchmarking its accuracy against HRP2-based RDTs, LM, and DBS-qPCR. A total of 672 whole blood samples were collected from 646 asymptomatic and 26 symptomatic individuals.

## RESULTS

### Malaria detection workflow overview

An overview of the adapted Dragonfly workflow is illustrated in **Fig. 1** and the entire standard operating procedure detailed in **Supplementary Methods.** On the front-end, an extraction method based on silica-coated superparamagnetic beads (TurboBeads, ProtonDx) and SmartLid technology was optimised to extract parasite DNA simultaneously from up to 12 whole blood samples in under 15 minutes, without using a centrifuge. Lyophilised colourimetric LAMP chemistry was then used for the rapid isothermal amplification of both pan-Plasmodium species and *Plasmodium falciparum* targets in a single reaction well, requiring only a simple low-cost (∼£100), and portable (160 x 110 x 130 mm, <1 kg), dry-bath heat block.^31,33,34^ Finally, results readout and interpretation were accomplished entirely visually, with a distinct colour change from pink (negative) to yellow (positive), avoiding expensive and bulky instrumentation required for fluorescent detection. Altogether, the entire sample-to-result workflow from EDTA-anticoagulated capillary blood was accomplished for up to 12 samples within 45 minutes by a single user.

**Figure 1.**
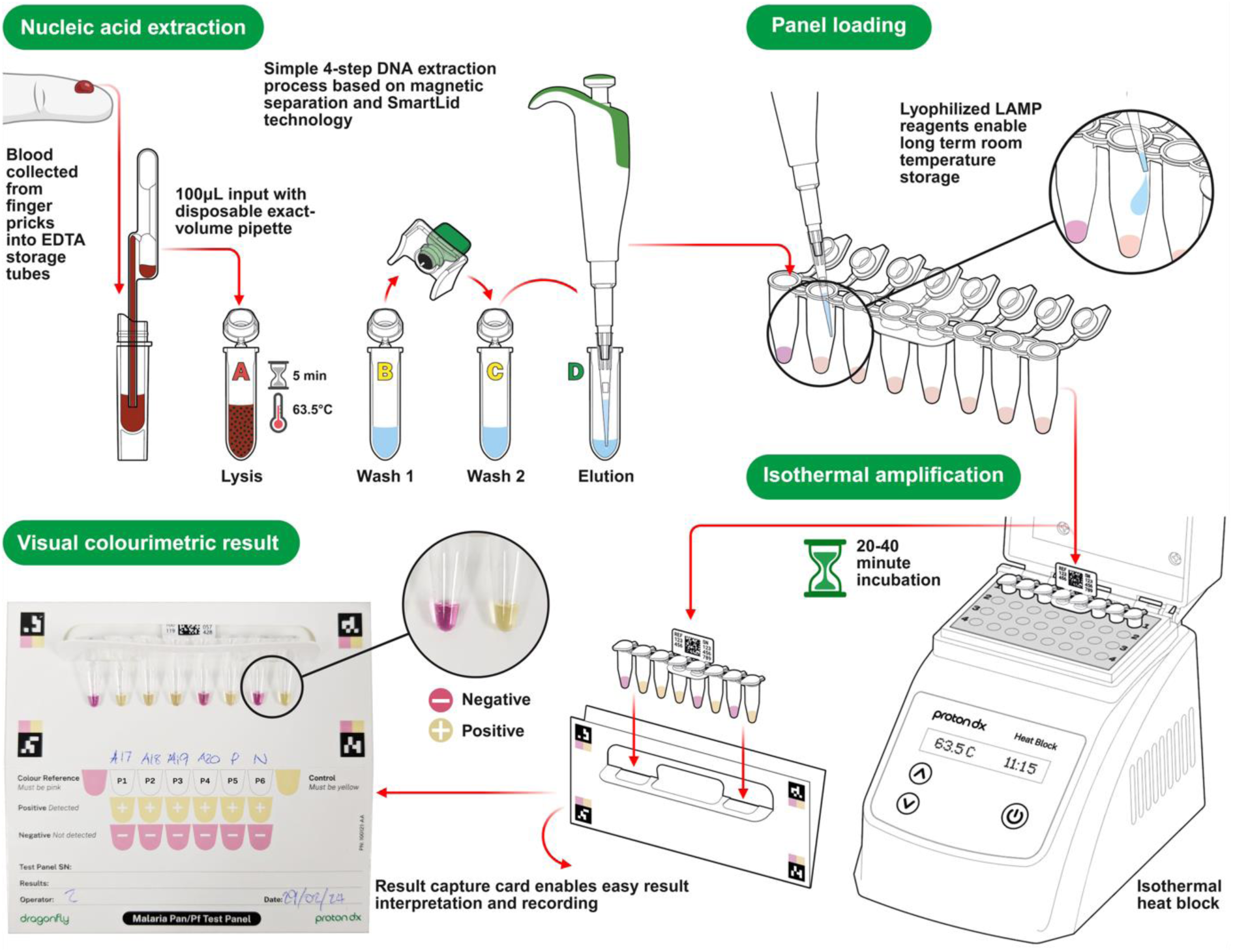
Schematic representation of our Pan/*Pf* malaria test workflow. The integrated system combines the SmartLid whole blood extraction process with LAMP-based isothermal amplification and colourimetric readout. The SmartLid extraction process for malaria detection from finger prick whole blood involves a four-step workflow (Lysis, Wash 1, Wash 2, and Elution) including a 5-minute heat-activated enzymatic incubation, and enables DNA purification and elution in under 10 minutes for a single sample. A total of 20 µL of eluted DNA is transferred using a fix-volume pipette into each reaction tube, followed by a maximum of 40-minute incubation at 63.5°C. Upon completion, LAMP results are qualitatively assessed by visually evaluating the colour change within the tubes, a pink colour indicating a negative result, while yellow a positive result. The validity of the test is confirmed by verifying that all controls exhibit the expected colour changes, as described below. Created in BioRender. Cavuto, M. (2025) https://BioRender.com/rwbh4tn.

### SmartLid blood DNA/RNA extraction kit

SmartLid-based nucleic acid extraction technology leverages a disposable lid with a removable magnetic key to quickly and easily transfer magnetic beads and attached nucleic acids through multiple buffers and steps in the extraction and purification process. After binding nucleic acids to the silica-coated magnetic beads (**Fig. 2a**), collection onto the lid is performed through multiple inversions of the tube with the magnetic key inserted (**Fig. 2b**). The SmartLid, along with the collected magnetic beads, can then be removed from the tube and transferred into the subsequent tube. Release of the magnetic beads into the new buffer is accomplished by removing the magnetic key and briefly shaking the tube. This entire process, transferring magnetic beads from one tube to another, is illustrated in **Fig. 2c**. Note that while the clear plastic (polypropylene) lid component of SmartLid is disposable, the green magnetic key is reusable, cutting down on plastic and rare-earth waste.

**Figure 2.**
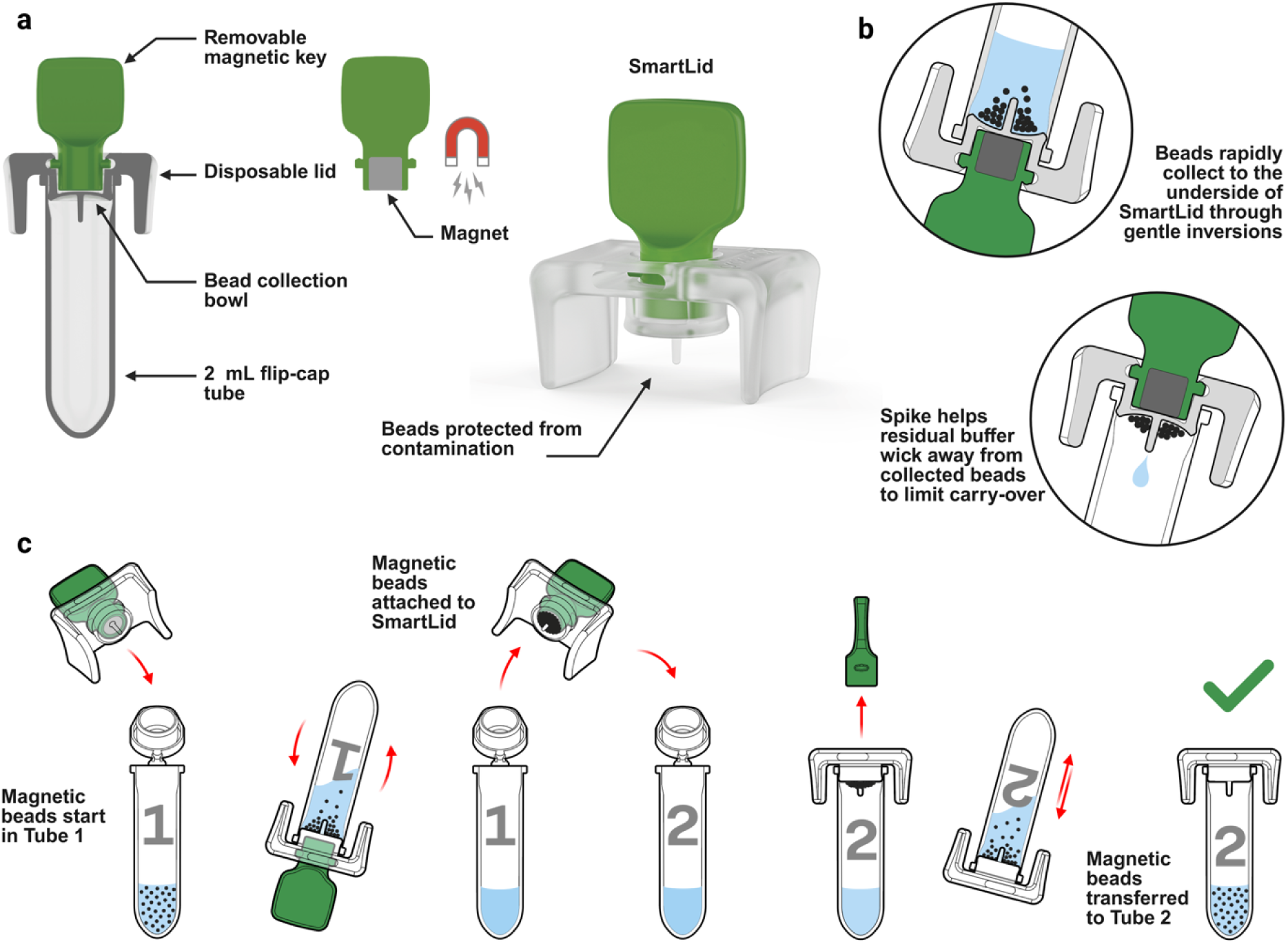
Illustrated overview of SmartLid technology. a) SmartLid is composed of two main components, a disposable clear plastic lid, designed to press-fit into most 2 mL flip-cap or screw-cap tubes, and a removable magnetic key, housing a 4 mm x 4 mm N42 neodymium magnet. b) Magnetic beads are collected onto SmartLid when the magnetic key is inserted, and the tube is inverted. A fluid wicking spike on the underside of the lid reduces buffer carry-over from tube to tube. c) The entire magnetic beads collection, transfer, and resuspension process is illustrated, which occurs multiple times throughout the SmartLid extraction process. Created in BioRender. Cavuto, M. (2025) https://BioRender.com/rwbh4tn.

The originally developed SmartLid protocol to extract viral DNA and RNA from swabs stored in guanidium thiocyanate-based buffer, eNAT transport/inactivation media (Copan, Italy), was adapted for this study to extract human and *Plasmodium* genomic DNA from 100 µL of EDTA-anticoagulated whole blood. Notably, a 5-minute heat-activated (65°C) enzymatic (proteinase K) lysis step was added to help break down the protein-rich sample matrix, along with an additional wash step to reduce contaminant carry-over into the elution. Finally, vortex mixing was utilised instead of manual shaking to agitate the magnetic beads in each extraction buffer, which is encouraged due to the higher viscosity and more inhabitant-rich sample matrix when compared to typical respiratory or skin swab eluent. For a single sample, the entire extraction process can be completed in approximately 10 minutes. All kits used for the study (SmartLid Blood DNA/RNA Extraction Kit) were custom produced in collaboration with ProtonDx Ltd (https://www.protondx.com/).

### SmartLid adaptation for medium-high throughput sample processing

The original SmartLid extraction method was developed for use at the POC and assumed processing only a single sample at a time with single use cardboard trays utilised both as the kit component packaging and as a workstation. For this study, as shown in **Fig. 3**, the SmartLid method was adapted for medium-high throughput sample processing by developing two key tools which together enabled the simultaneous processing of up to 12 samples at a time by a single user. First, a 3D printed (X1 Carbon with AMS, Bambu Labs) tube rack was created, with 12 columns (one for each sample) of four rows (one for each step in the sample extraction process), labelled 1-12 and A-D for the columns and rows, respectively **(Fig. 3a).** The spacing of each column and row was optimised to enable easy transfer of SmartLids from tube to tube within a column without obstruction from the open flip-cap lids. Users were also encouraged to write the column number on top of each SmartLid to further reduce the likelihood of accidentally mixing up two samples. Next, a multi-tube vortex tool was developed to conveniently hold up to 12 sample tubes (with attached SmartLids), enabling simultaneous mixing **(Fig. 3b-f)**. The central column on the underside of the vortex tool is depressed into the centre of the vortex mixer, while a screw-on lid with a handle clamp down on the top of each SmartLid. Finally, each tube location in the tool is numbered to allow easy correlation with the number on each SmartLid and/or column number in the tube rack. Combined, these two new tools enabled 12 whole blood samples to be extracted in parallel in under 15 minutes by a single user.

**Figure 3.**
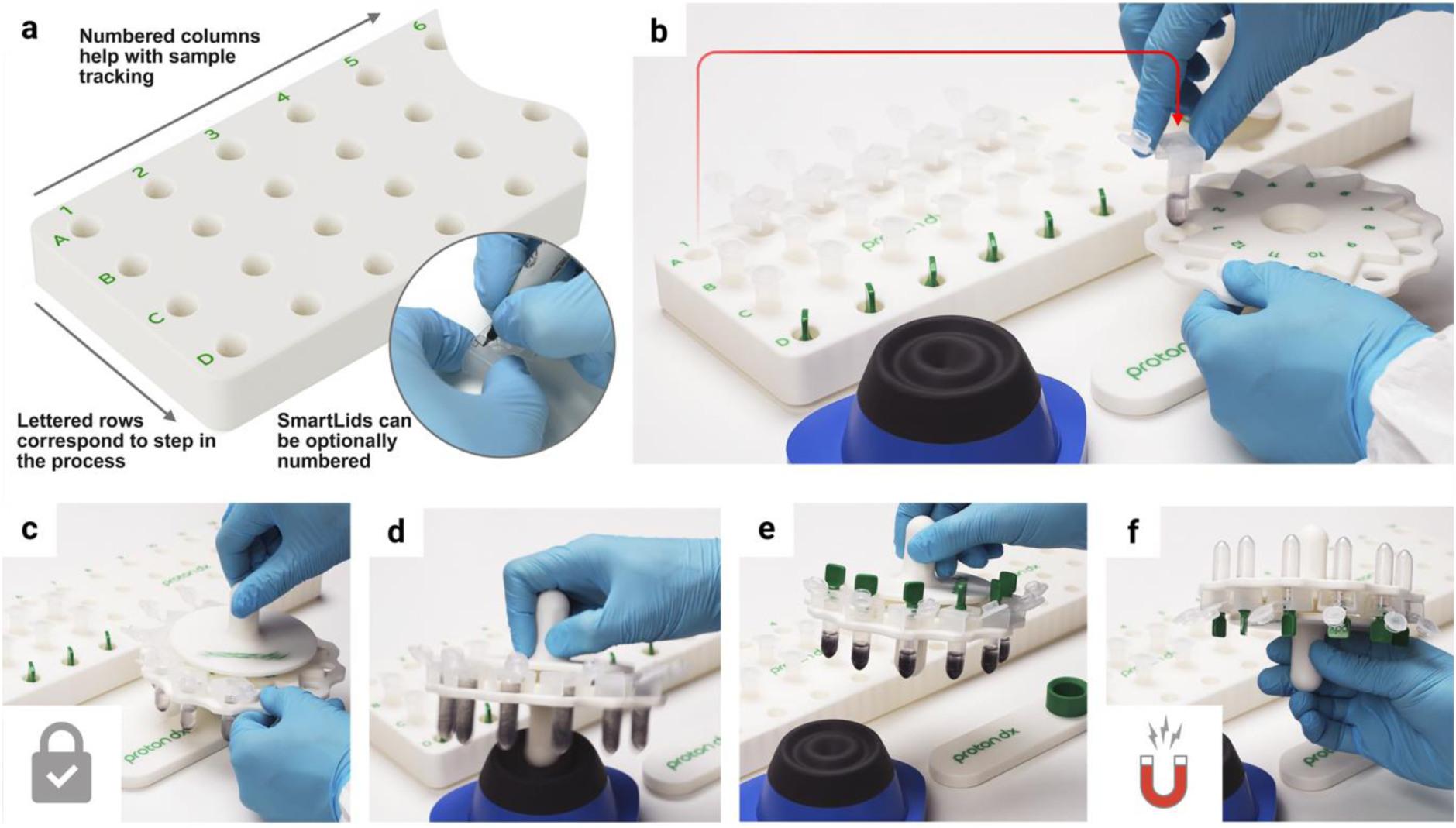
Summary of SmartLid accessories to enable medium-high throughput sample processing. a) The SmartLid Rack, with numbered columns (1-12) to identify the sample and lettered rows (A-D) for each step in the extraction process. b) A tube being transferred from the SmartLid Rack into the SmartLid Vortex Tool by a user performing six simultaneous extractions. c) A screw-on plate locks all tubes and maintains all SmartLids in place while also providing a handle. d) All samples are mixed simultaneously and equally by depressing the top of the vortex mixer with the central underneath column of the tool. e) All tubes are fully mixed and magnetic keys are inserted into each SmartLid. f) All magnetic beads are collected simultaneously through inverting the Vortex Tool. Created in BioRender. Cavuto, M. (2025) https://BioRender.com/rwbh4tn.

### Multi-patient malaria Pan/*Pf* test panel design

The Pan/*Pf* malaria test panel presented here was adapted from the single-patient, multi-pathogen format to a format designed to test multiple patients for a single pathogen in order to increase throughput and reduce cost per patient.^32^ Each flip-cap tube within the 8-tube strip panel contained lyophilized colourimetric LAMP reagents which are stable at room temperature for extended periods and provide a clear visual colour change from pink to yellow to indicate a positive result. The tandem Pan/*Pf* assay was designed to target both Pan-malaria DNA sequences, conserved across multiple *Plasmodium* species responsible for malaria, and *Plasmodium falciparum*-specific sequences. A comprehensive list of the previously published primer sequences used in this study is provided in **Supplementary Table 1**.^20,35^

In addition to six Pan/*Pf* target reactions, allowing for the simultaneous screening of six individuals, two additional control reactions are included in the 8-tube strip panel to ensure validity of results. First, as the reaction utilises an unbuffered LAMP system and pH sensitive dye to detect amplification, a colour reference control reaction was included, which omits polymerase enzymes to prevent amplification. This reaction will always remain pink with the exact shade of pink varying based on the starting pH of the eluted nucleic acids. Depending on the sample type (*e.g.* capillary blood from finger pricks *versus* dried blood spot eluates), this starting pH and subsequent colour can vary slightly. Therefore, this reaction provided a reference colour against which all other reaction outcomes were compared. An internal control reaction was also included, targeting an exogenous DNA template lyophilized with the rest of the reaction reagents, which amplified if the correct incubation temperature was reached, the reaction was incubated for sufficient time, and reagents were not damaged in storage.

Reaction positions in the 8-tube strip, from left to right, were as follows: colour reference control (tube 1), six independent Pan/*Pf* reactions (tubes 2–7), and the internal control (tube 8). Each reaction was reconstituted with 20 µL of sample eluate. Isothermal amplification was conducted using a portable dry bath heat block (ProtonDx, UK) set at 63.5 °C. Finally, while the heat block was powered by mains electricity in this study, it is also compatible with portable batteries, standard 12-volt supplies, or solar panels. It requires less than 20W of continuous power to maintain the set temperature, supporting its suitability for decentralized testing in resource-limited settings.

### Assessment of analytical specificity and incubation time

To assess the risk of false positive, the analytical specificity of our test was evaluated at different incubation time points up to 60 minutes. At 50 minutes, the test demonstrated a specificity of 98.3% (95% CI: 96.0–100%), with 2 false positive results out of 120 negative samples tested, as shown in **Supplementary Table 2**. A maximum incubation time of 40 minutes was required to confirm negative results, although most positive reactions turned yellow within 20 minutes.

### Comparison of analytical sensitivity using in vitro cultured ring stages (3D7 strain)

Using serial dilutions of 3D7 parasite cultures, both LAMP platforms, Dragonfly (input volume 100 µL) and Alethia® (input volume 50 µL), consistently detected all replicates down to a parasitaemia of 0.9 parasites/μL, outperforming DBS-qPCR (input equivalent to 3 discs of 3 mm diameter), which showed decreased sensitivity below 3.8 parasites/μL. WB-qPCR (input volume 100 µL) demonstrated the highest analytical sensitivity among the four molecular-based methods **(Fig. 4a).** The LODs (parasites/μL) estimated by probit analysis were 2.9 [95% CI: 1.8-4.8], 0.7 [95% CI: 0.3-1.3], 0.6 [95% CI: 0.3-1.4], and 0.4 [95% CI: 0.2-0.6] for DBS-qPCR, Alethia®, Dragonfly, and WB-qPCR, respectively (**Fig. 4b**).

**Figure 4.**
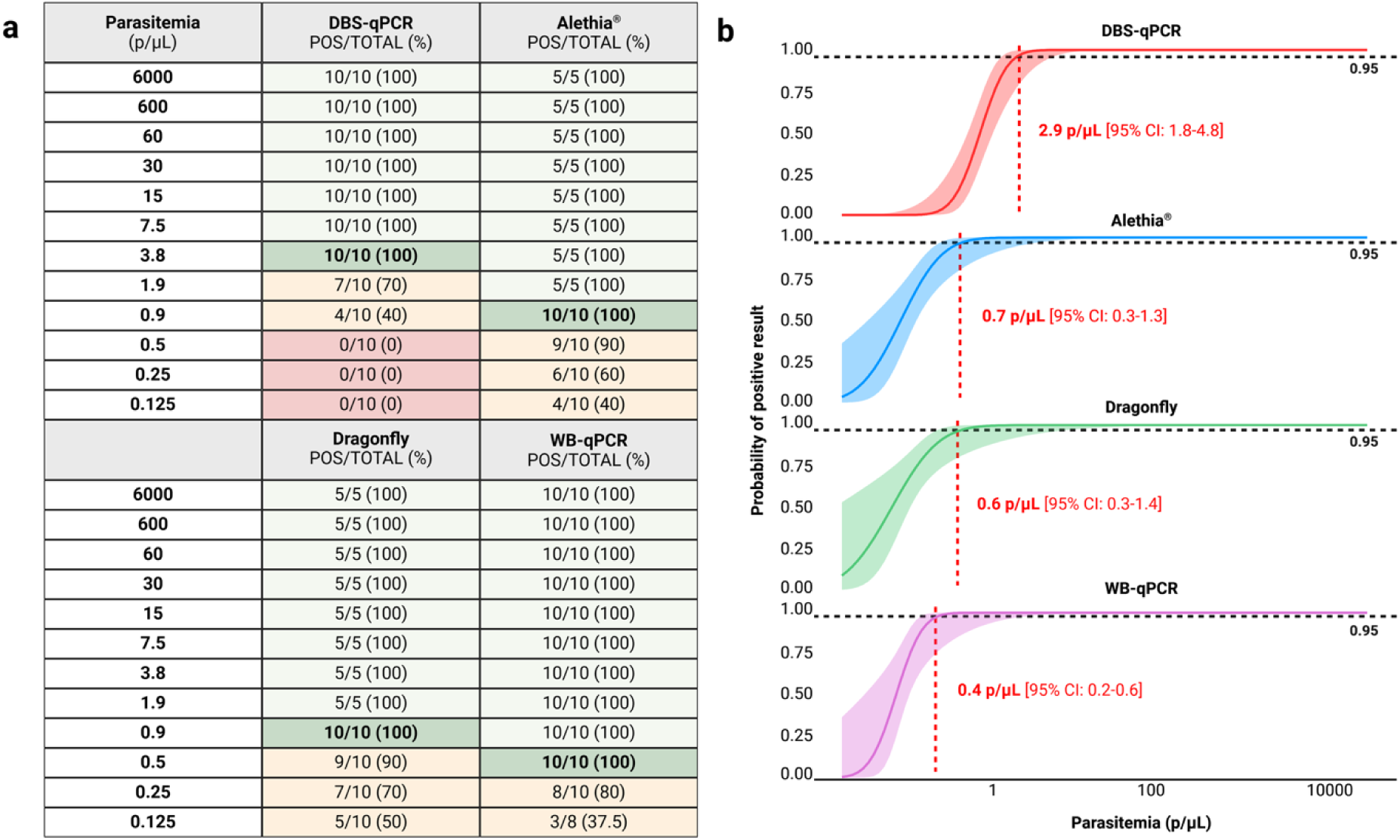
Comparison of analytical sensitivity of malaria detection using spiked whole blood across Dragonfly, Alethia®, DBS-qPCR, and WB-qPCR. Experiments were conducted in collaboration with the London School of Hygiene and Tropical Medicine (LSHTM) using spiked EDTA-blood with ring-stage *Plasmodium falciparum* 3D7 strain. Results are shown in terms of (a) number and percentage of successfully detected replicates at each spike concentration as well as (b) the resulting empirically determined LOD through probit analysis. The LOD is defined as the parasite density at which the probability of a positive result is ≥95%. Created in BioRender. Cavuto, M. (2025) https://BioRender.com/rwbh4tn.

### Validation of the Dragonfly platform against RDT and light microscopy using DBS-qPCR as the gold standard

First, a total of 50 capillary blood specimens from febrile malaria patients, purposively selected as positive controls based on concordant *Plasmodium* detection by LM, RDT, and DBS-qPCR, were also tested using the Dragonfly platform. All 50 samples tested positive, confirming the compatibility of our method with real, field-collected capillary blood samples prior to evaluation on unlabelled community survey samples. Next, 672 capillary blood samples collected at the community-level in The Gambia and Burkina Faso were used to evaluate the performance of the Dragonfly platform against DBS-qPCR as the reference method. All samples were also assessed using expert LM and RDTs to benchmark the performance of our method against the two standard diagnostic approaches for malaria. Of the 672 samples, 27.1% (146/672) were positive for *P. falciparum* by DBS-qPCR. These positive samples represented a broad range of parasite densities, including both microscopically detectable (n=103) and submicroscopic infections (n=43). A breakdown of the sample categories is presented in **Fig. 5**. Detailed results obtained using Dragonfly, Alethia®, DBS-qPCR, and WB-qPCR for each of the 50 positive controls from confirmed malaria patients, as well as the 672 capillary blood samples, are provided in the **Supplementary Data**.

**Figure 5.**
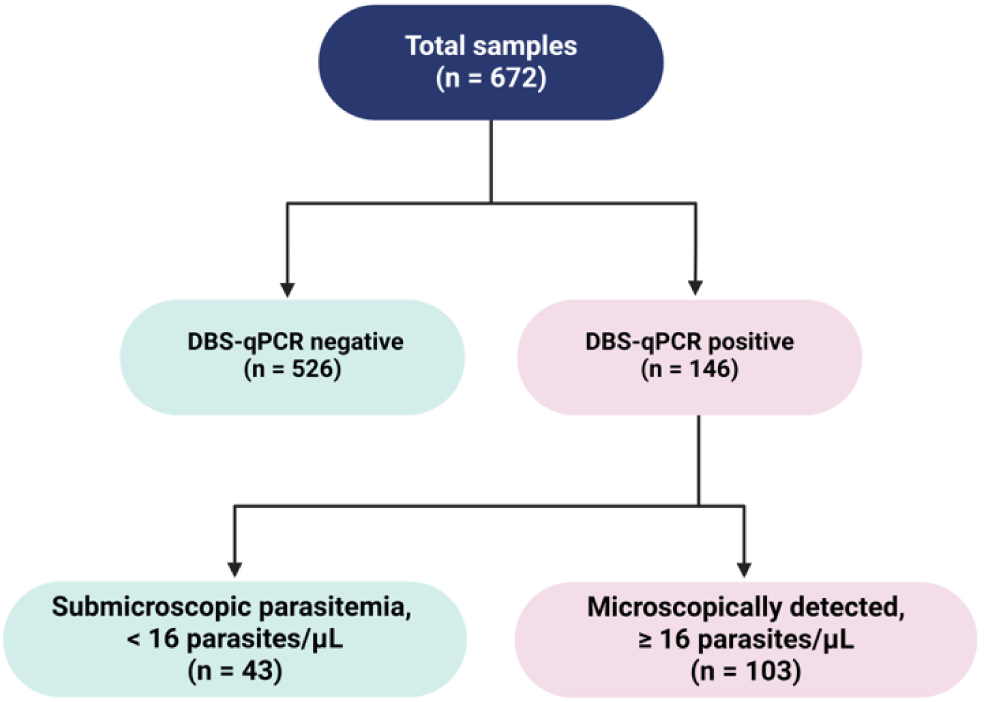
Clinical samples selection to evaluate the performance of Dragonfly Pan/*Pf* malaria platform. Submicroscopic parasitaemia is defined as a parasite density of < 16 parasites/μL, corresponding to the theoretical LOD for an expert microscopist, based on the ability to detect one asexual parasite among 500 leukocytes, assuming a white blood cell count of 8,000 leukocytes/μL. Created in BioRender. Cavuto, M. (2025) https://BioRender.com/rwbh4tn.

As depicted in **Fig. 6**, considering the 672 samples, the overall sensitivity and specificity of our method against DBS-qPCR was 95.2% [95% CI: 90.4–98.1] and 96.8% [95% CI: 94.9–98.0], respectively. Comparatively, the Dragonfly method demonstrated a higher sensitivity than both RDT (50.7% [95% CI: 42.3-59.0]) and expert LM (70.5% [95% CI: 62.4-77.8]). All three methods achieved specificities above 96%, with expert LM recording the lowest false-positive rate.

**Figure 6.**
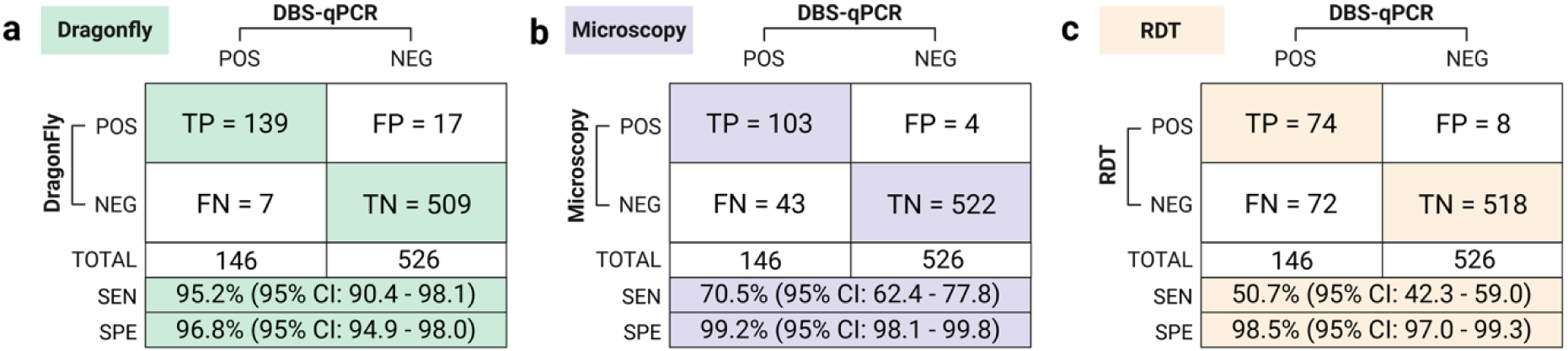
Comparison of the clinical performance of Dragonfly, RDT, and LM using whole blood finger prick samples, with DBS-qPCR as the gold-standard comparator. For each group, the number of true positives (TP), total positive cases, sensitivity rate with 95% CI, number of true negatives (TN), total negative cases, and specificity rate with 95% CI are provided. FP= false positive, FN=false negative. Created in BioRender. Cavuto, M. (2025) https://BioRender.com/rwbh4tn.

When considering samples from asymptomatic individuals only (N=646, including 137 malaria positive and 509 negative samples determined by DBS-qPCR) the sensitivity gap remained similar, with Dragonfly, LM, and RDTs detecting 94.9%, 70.1%, and 49.6% of positive samples, respectively. Confusion matrices for this subset of samples, as well as the smaller subset of symptomatic cases, are provided as **Supplementary Fig. 1**.

As summarised in **Fig. 7**, the 146 DBS-qPCR-positive specimens were stratified into four parasite-density groups: <16 parasites/µL (n = 43), 16–100 parasites/µL (n = 28), 100–200 parasites/µL (n = 13), and >200 parasites/µL (n = 62). Across the three lower-density categories Dragonfly markedly outperformed RDT. Dragonfly detected 41 of 43 specimens in the <16 parasites/µL group (95.3 %), 25 of 28 specimens in the 16–100 parasites/µL group (89.3 %), and all 13 specimens in the 100–200 parasites/µL group (100 %). In contrast, the RDT detected 2 of 43 specimens (4.7 %), 9 of 28 specimens (32.1 %), and 8 of 13 specimens (61.5 %) in the corresponding groups, respectively. Dragonfly also demonstrated significantly higher sensitivity than expert LM for sub-microscopic parasitaemia (<16 parasites/µL); however, no significant differences (p > 0.05) were observed between Dragonfly and expert LM in the 16–100 and 100–200 parasites/µL groups. When parasite density exceeded 200 parasites/µL, all three methods showed comparable performance (p > 0.05).

**Figure 7.**
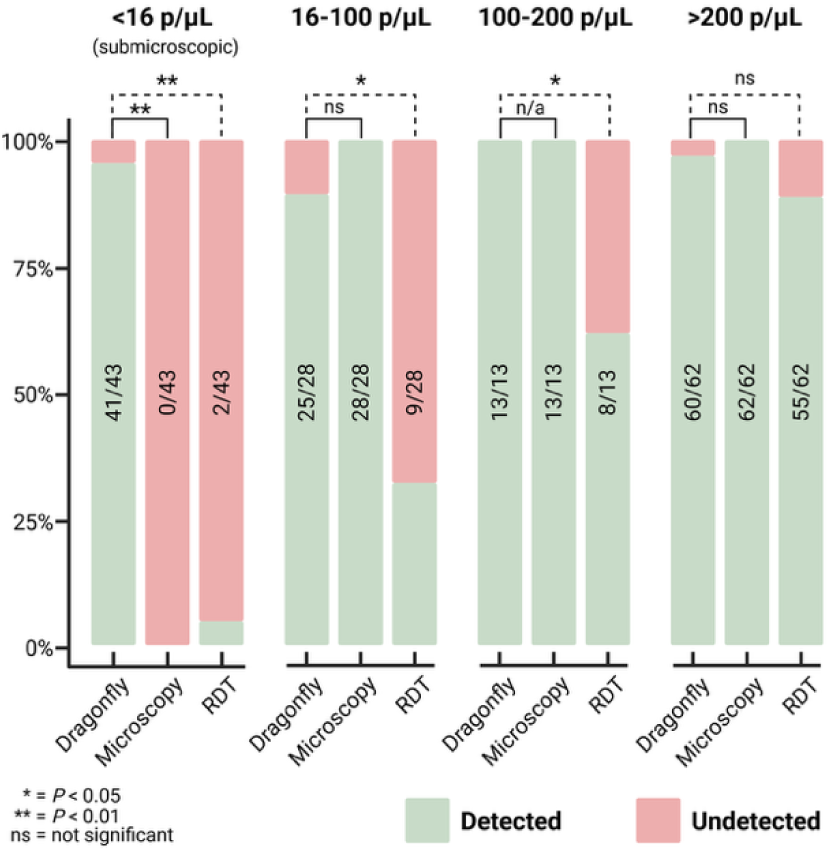
Detection of DBS-qPCR positive samples by Dragonfly, RDT, and expert LM, stratified by four parasite density categories: <16, 16–100, 100–200, and >200 parasites/µL. Dragonfly correctly identified 95.3% of submicroscopic infections, 41 out of 43 samples detected with <16 parasites/µL, significantly outperforming both expert LM and RDT. Dragonfly also significantly outperformed RDT at densities ranging from 16–200 parasites/µL, with no significant difference observed between Dragonfly and expert LM in this range. At parasite densities >200 parasites/µL, no statistically significant differences were observed among the three methods (p > 0.05). Created in BioRender. Cavuto, M. (2025) https://BioRender.com/rwbh4tn.

## DISCUSSION

Through collaborative efforts between UK and West African institutions, this study presented a near-POC colourimetric LAMP-based extracted molecular solution to achieving accurate detection of sub-microscopic *Plasmodium* infections from whole blood at the community level in Sub-Saharan Africa. Our approach demonstrated high analytical performance, achieving an LOD of 0.6. parasites/µL [95% CI: 0.3-1.4] with spiked samples. Field evaluation demonstrated sensitivity and specificity of 95.2% [95CI: 90.4-98.1%] and 96.8% [95CI: 94.9-98.0%] from individuals enrolled at community level, most of them (96%) asymptomatic. This high diagnostic accuracy, along with the ability to detect 95.3% of the submicroscopic infections (<16 parasites/µL), most of which were missed by both RDT and LM, suggests that the approach may be considered a valuable tool for community-based ADIs in malaria-endemic regions. Moreover, all readings in our study were performed by expert microscopists to ensure accurate identification of parasites, especially for low parasite density infections. This rigorous approach likely contributed to the higher sensitivity by LM observed in our study compared to routine clinical practice, where blood slide may be read by less experienced technicians.

Since the introduction of LAMP technology for malaria detection, more than 26 LAMP assays have been developed and evaluated, demonstrating an estimated pooled sensitivity of 97.1% [95CI: 95.7-98.0%] as reported in a previous meta-analysis study including both symptomatic and asymptomatic individuals.^39^ However, to date only two LAMP-based diagnostic tests (Loopamp^TM^ Malaria and Alethia® Malaria) are currently commercialised for malaria, suggesting that high technical performance alone is insufficient to ensure the sustainable deployment of a diagnostic test in its intended target settings.^19,24^. Performance studies conducted in both health facilities and community settings demonstrated varying sensitivities of these two commercial options, ranging from 97.2% to 40.8% for symptomatic and asymptomatic cases, respectively.^40–44^ In our study, Alethia® demonstrated comparable high analytical performance, which combined with its user-friendliness likely contributes to its widespread adoption in high-income countries to guide malaria diagnosis in returning travellers.^28–30^ However, its deployment in rural African settings, especially for large-scale community-based malaria screening interventions is constrained by its high cost and the limited capacity of the Alethia incubator accommodating only 10 samples per run. In contrast, Dragonfly combines high diagnostic accuracy with several key advantages for field deployment in resource constrained settings, meeting all the essential (and many of the desirable) technical and health systems criteria outlined in the malERA Target Product Profiles for diagnostics intended for malaria screening and surveillance, as detailed in **Supplementary Table 3.**^17^ These specifications include, ease of sample collection, high sensitivity and specificity, rapid turnaround time, ease of use and portability. For example, the SmartLid DNA extraction method is compatible with capillary finger prick whole blood, enabling field sampling using well accepted procedures (finger pricks blood samples) in routine healthcare and malaria testing across Sub-Saharan Africa. Furthermore, the SmartLid technology and highly optimized protocol ensures quality nucleic acid extraction and purification in a fraction of the time (< 15 minutes for up to 12 samples at a time) of gold-standard PCR methods which can take well over an hour and rely on bulky and expensive centrifuges. On the detection side, the lyophilised isothermal colourimetric LAMP chemistry forgoes the cost and bulk of thermocyclers and devices that rely on LED illuminators or fluorescent detectors to determine the result. Altogether, 12 whole blood samples can be extracted and amplified from start to finish in as little as 35 minutes for high parasitaemia samples, relying only on a vortex mixer and two low-cost isothermal heat blocks for powered equipment, and room temperature storage for all consumables. Further comparison of the characteristics of the Dragonfly platform with the two commercial Malaria LAMP technology is shown in **Supplementary Table 4**.

ADIs such as Mass-Testing-and-Treatment (MTAT) and Focused Testing and Treatment (FTAT) are currently not recommended by WHO due to their limited or negligible impact on malaria prevalence and incidence of clinical malaria.^36,37^ However, such recommendations are primarily based on intervention trials that used RDT and/or LM for malaria diagnosis.^36,37^ Recent modelling studies suggested that deploying a diagnostic test with sufficiently high sensitivity, such as one that reduces the limit of detection below 200 parasites/µL, can accelerate malaria elimination, provided a high coverage of sufficient duration is achieved, and the treatment is efficacious.^8,9,38^ It has been further shown that by reducing the LOD below 2 parasites/µL, MTAT strategies could substantially increase the identification of sub-microscopic cases, leading to a reduction in *Plasmodium falciparum* PCR-based prevalence and a decrease in the required number of intervention rounds.^9^

Since our platform is still in a prototype development stage, a comprehensive cost analysis of the final manufactured version is currently unavailable but will be a significant factor in the assay’s suitability for deployment in sub-Saharan Africa. However, a preliminary cost analysis is provided in **Supplementary Table 5**, based on prototype quantities and estimations at low- and medium-scales, and is compared to the current market costs of the Alethia® Malaria Test and associated required instrument. Another limitation of our study is that Dragonfly malaria testing was performed in standard laboratories in The Gambia and Burkina Faso which do not reflect the real-life conditions of field sampling and testing. Therefore, future studies will assess the robustness of the device in more decentralized environments such as community settings and gather detailed insights into user experiences and device usability in the field. In preparation for this investigation, the SmartLid extraction method described here is being adapted into the true-POC single-use format of the previously presented Dragonfly sample-to-result platform,^32^ with all reagents and buffers pre-aliquoted and laboratory micro-pipettes replaced with disposable exact volume pipettes. Finally, although the technology development and adaptation were performed in laboratories in London through an active collaboration with African Institutions, future initiatives should focus on extending the concept of transferability to local development and production in Africa. This approach would facilitate the sustainable manufacturing and distribution of the diagnostic platform, thereby ensuring its availability in malaria-endemic regions.

In addition to demonstrating the strong potential of the SmartLid extraction and Dragonfly colourimetric LAMP technologies towards malaria elimination strategies, this work highlighted the rapid adaptability of the combined approach and potential for assay transfer, which should be a key criterion for selecting molecular diagnostic tests in Sub-Saharan Africa.^45^ Our approach’s significant use of off-the-shelf consumables and flexibility enabled the rapid prototyping and deployment of the presented multi-patient Pan*/Pf* malaria test from capillary finger prick blood samples. Finally, the potential for future digital and cloud integration of the Dragonfly platform, as explored in previous works demonstrating solutions for viral respiratory and skin infection diagnostics, aligns with the growing need for connected diagnostics solutions in Africa.^32,46^

## ONLINE METHODS

### Comparison of analytical sensitivity using in vitro cultured ring stages (3D7 strain)

The analytical sensitivity of Dragonfly was evaluated in comparison with Alethia®, DBS-qPCR and WB-qPCR. The comparison was performed using serial dilutions of ring-stage *Plasmodium falciparum* (3D7 strain) parasite culture at the malaria parasitology lab of the LSHTM in London. Parasites were cultured in-vitro and synchronized using a magnetic separation procedure.^47^ To minimise the occurrence of red blood cells (RBCs) infected by multiple parasites, cultures were maintained under gentle agitation (60RPM), resulting in 85% of singly infected RBC among the total infected cells (**Supplementary Fig. 2**). The final parasitaemia of the culture, measured at 5.8% of RBCs, was confirmed by expert LM. A Plasmodium-negative blood sample (tested by WB-qPCR) was then spiked to generate the initial infected sample, yielding a parasitaemia of 6,000 parasites/µL. Serial dilutions were subsequently performed using the same negative whole blood sample to generate samples with decreasing concentrations, calculated based on the dilution factor applied. Each dilution point was tested multiple times across the four molecular-based methods. The concentrations of each dilution point, as well as the number of replicates per method, are summarized in **Fig. 4**.

Alethia® is a commercially available LAMP-based technology that utilises a *Plasmodium* genus-specific assay. The system consists of an initial DNA extraction process using a passive gel filtration column, followed by an amplification step with lyophilised reagents in a dedicated LAMP incubator. The test provides on a LCD screen a qualitative result (positive or negative), which is automatically interpreted by a reader integrated within the incubator. As per the manufacturer’s guidelines, 50μL of whole blood was used as the input volume in our evaluation.

DNA was extracted from DBS (3 discs of 3 mm) and whole blood samples (100 µL) using the QIAamp DNA Mini Kit according to the manufacturer’s instructions. DNA was eluted in AE Buffer with a final volume of 100 µL for DBS and 200 µL for whole blood samples. The *Plasmodium falciparum*-specific PCR assay applied to both sample types has been previously described.^48^ The PCR reaction volume was 20 µL, comprising 5 µL of DNA extract, 10 µL of GoTaq qPCR Master Mix 2X, 2 µL of *P. falciparum* primer/probe mix (F/R/P] 10X, and 3 µL of PCR grade water. Amplification was performed on a LightCycler® using the following cycling conditions: an initial denaturation at 95 °C for 2 minutes, followed by 45 cycles of denaturation at 95 °C for 15 seconds and annealing/extension at 60 °C for 1 minute.

### Dragonfly malaria field validation against RDT and light microscopy *versus* DBS-qPCR

The compatibility of the adapted Dragonfly method with finger prick clinical samples was assessed using 50 blood specimens purposively selected based on their positivity for *Plasmodium* by RDT, LM and DBS-qPCR. These specimens were obtained from febrile patients attending rural health facilities in the Central Region in Burkina Faso. Parasite densities, determined by expert LM, ranged from 149 to 87,500 parasites/µL with a median [IQR] of 714 parasites/µL [95% CI: 128-5,688 parasites/µL].

A total of 672 blood specimens were tested to evaluate the performance of the presented method compared to RDT and LM using DBS-qPCR as gold standard. Blood specimens were collected from individuals enrolled in a community-based survey in two malaria endemic sites, *i.e.,* The Central Region in Burkina Faso and The Upper River Region in The Gambia, characterised by a predominance of *P. falciparum.* Baseline characteristics of study participants are summarized in **Table 1** and show a slight predominance of females (57%) and a fair representation of all age categories. Most participants (96%) were asymptomatic at the time of sample collection.

**Table 1.** Characteristics of study Participants.

| Characteristics | n (%) |
| --- | --- |
| **Sex** |  |
| Male | 289 (43.0) |
| Female | 383(57.0) |
| **Age groups in years** |  |
| 0.5–4 | 78 (11.6) |
| 5–14 | 198 (29.5) |
| 15–29 | 126 (18.7) |
| 30–59 | 188 (28.0) |
| ≥60 | 82 (12.2) |
| **Symptomatic status** |  |
| Symptomatic | 26 (3.9) |
| Asymptomatic | 646 (96.1) |
| **Sample collection sites** |  |
| The Gambia | 281 (41.8) |
| Burkina Faso | 391 (58.2) |

All malaria tests were performed using capillary blood collected by finger prick. RDT testing was performed on-site, either in a health facility (50 blood specimens used for the compatibility assessment) or at community level (672 blood specimens used for the diagnostic performance evaluation). LM readings were performed on thick smears. Clinical blood specimens used for the Dragonfly method collected into 200 μL EDTA microtainers were stored between 3-5 °C if they were tested the same day as sample collection, or otherwise stored at –20 °C until testing. Blood spots from the same finger prick were collected on DBS card and used for qPCR analysis.

### Malaria confirmation by DBS-qPCR

DBS-qPCR was selected as the gold standard method for evaluating the performance of the developed platform. The DBS-qPCR testing was conducted at the MRCG following a standardized protocol.^49^ The gold standard method consisted of capillary blood spotted onto Whatmann filter paper followed by a qPCR detection using a TaqMan assay targeting the *var* gene acidic terminal sequence of *Plasmodium falciparum*. The number of copies of the *var* gene is approximately 59 per genome. Genomic DNA extraction was performed using the QIAamp 96 DNA QIAcube HT Kit, in accordance with the manufacturer’s instructions. Three discs of 3mm diameter were punched out of the DBS and the DNA extracts were eluted in a volume of 80 µL of AE Buffer. DNA amplification was conducted using the Bio-Rad CFX96 real-time PCR machine. Each reaction utilized 5 µL of sample DNA. The VarATS qPCR master mix consisted of 1 µL of PCR-grade water, 10 µL of 2X TaqMan Master Mix, 1.6 µL of 10 µM *Var* forward primer, 1.6 µL of 10 µM *Var* reverse primer, and 0.8 µL of 10 µM Var probe. The cycling conditions were: 1 cycle at 50 °C for 2 min and 1 cycle at 95 °C for 10 min followed by 45 cycles at 95 °C and 55 °C for respectively, 15sec and 1min. If the result was positive for either PCR or Dragonfly alone, the PCR was repeated in duplicates starting from a new DNA extraction. If at least one of the repeats was positive, the sample was considered positive for *Plasmodium falciparum*.

### Light microscopy

The thick blood smear was prepared from fresh capillary blood and air dried. Giemsa staining was performed with a 3% solution for 30 minutes. All slides were independently read by two qualified microscopists. Parasite density was determined by counting the number of asexual parasites per 500 leukocytes on the thick smear, assuming a leukocyte count of 8,000/µl, and using 100x magnification with optical LM. In case of discrepancy between the two readers such as one reporting negative and the other positive for malaria or a density difference of > 50%, or different species reported, a third expert microscopist would read the slide as well. The reported parasite density was the geometric mean of the two readers’ results or that of the two closest readings if a third reading was done.

### Rapid-diagnostic tests

*Plasmodium falciparum* infection status of all 672 community-collected samples was assessed using the SD BIOLINE Malaria *P. falciparum* Ag Test™ (Abott), an *HRP2*-based immunolateral flow assay accepted on the WHO list of prequalified *in vitro* diagnostics.^50,51^

### The Dragonfly Pan/*Pf* malaria test workflow

Extractions were performed manually using the SmartLid sample preparation in bulk reagents format allowing for efficient processing of multiple samples simultaneously. A total of 100 µL of capillary blood sample was used as input for each extraction. Purified samples were eluted in a total elution volume of 50 µL. LAMP reactions were performed by adding 20 µL of the eluted sample into each tube of the colorimetric LyoLAMP Pan/*Pf* Test Panels (ProtonDx). The Pan-Plasmodium (Pan18s) *and P. falciparum (*PfK13, Pf mtDNA) assays’s genes and primer sequences used in this study are listed in **Supplementary Table 1.** Amplification was conducted in the portable thermal block (ProtonDx) for 40 min at 63.5 °C. Immediately following amplification, results were visually assessed by the technician based on the colour change in each tube, with a change from pink (negative) to yellow (yellow) in case of *P. falciparum* infection.

All testing was carried out in standard molecular biology laboratories at MRCG Unit The Gambia and Clinical Research Unit Nanoro, Burkina Faso. All steps, from DNA extraction to result-read-out, were conducted in a single workspace without the need for a laboratory hood. Prior to testing, a half-day training program was conducted for users including both theoretical instructions and hands-on practices using 3D7-infected whole blood and negative controls.

### Data Analysis

All samples were fully anonymized prior to analysis. Data from the specificity and analytical sensitivity experiments are provided in **Supplementary Data**. Socio-demographics, DBS-qPCR, RDT, LM and Dragonfly data of the total clinical samples (n=672) were merged into a unique database and statistical analyses conducted using Stata 18 (StataCorp, College Station, TX, USA) and R (R Core Team, Vienna, Austria) (**Supplementary Data**). Summary statistics were performed using the median value and IQR for continuous variables, and proportions and 95%CIs for categorical variables. Sensitivity and specificity with their 95% confidence intervals were calculated for Dragonfly, LM and RDT using DBS-qPCR as the gold standard according to formulas shown in **Supplementary Table 6**. The LOD was estimated using probit analysis defining LOD as the concentration with a 95% probability of obtaining a positive result.^52^ To assess the differences between groups, McNemar’s Test was employed. A p-value < 0.05 was considered statistically significant.

### Ethics

Collection of samples during community-based surveys in The Upper River Region in The Gambia was approved by the LSHTM Ethics Committee and The Gambia Government/MRC Joint Ethics Committee (Ref. 29611). In Burkina Faso, sample collection from symptomatic patients enrolled at health facilities in the Central West Region received approval from The Comite d’Ethique pour la Recherche en Sante-Burkina Faso (ref: DELIBERATION N°2021-04-084) and the Comité d’Éthique Institutionnel pour la Recherche en Sciences de la Santé of IRSS (N/Réf. A07-2021/CEIRES). The approval for collecting samples from participants enrolled at community level was granted by The Comite d’Ethique pour la Recherche en Sante-Burkina Faso (ref: DELIBERATION N° 2024-0361). Written informed consent was obtained from all research participants and/or guardians before recruitment.

## ACKNOWLEDGEMENTS

This work was supported by the Department of Health and Social Care-funded Centre for Antimicrobial Optimisation (CAMO) at Imperial College London; the London School of Hygiene and Tropical Medicine (LSHTM), the Wellcome Trust CAMO-Net programme [226691/Z/22/Z]; the Wellcome Trust Innovator Award [215688/Z/19/Z]; and the Imperial College Research Fellowship to KMC [WDPI.G09074]. In addition, this work was funded by the NIHR [NIHR134694] using UK aid from the UK Government to support global health research. Infrastructure support for this research was funded by the NIHR Imperial Biomedical Research Centre (BRC). The views expressed in this publication are those of the authors and not necessarily those of the NIHR or the UK Department of Health and Social Care. P.G. and J.R.M. are affiliated with the NIHR Health Protection Research Unit (HPRU) in Healthcare Associated Infections and Antimicrobial Resistance at Imperial College London in partnership with the UK Health Security Agency, in collaboration with Imperial Healthcare Partners, the University of Cambridge and the University of Warwick.

## AUTHOR CONTRIBUTIONS

D.R.R., I.P. and M.L.C. contributed equally to the work. Study concept and design: D.R.R., I.P., A.E., U.D.A., H.T., A.C., and J.R.M. Acquisition, analysis, or interpretation of data: D.R.R., I.P., M.L.C., F.K., M.C., D.Y.S., E.Q., K.M.C., M.O.N., S.C., B.D., L.B.S., P.G., M.B., H.T., A.J.C., A.E., U.D.A., and J.R.M. Drafting the manuscript: D.R.R., I.P., M.L.C., J.R.M. Critical revision of the manuscript: D.R.R., I.P., M.L.C., F.K., M.C., D.Y.S., E.Q., K.M.C., M.O.N., S.C., B.D., L.B.S., P.G., M.B., H.T., A.J.C., A.E., U.D.A., and J.R.M. The manuscript was written through the contributions of all authors. All authors have given approval to the final version of the manuscript.

## DATA AVAILABILITY

All data supporting the findings of this study are available within the article and its supplementary files. Any additional requests for information can be directed to, and will be fulfilled by, the corresponding authors. Source data are provided with this paper.

