## Supplementary Information for "High-Sensitivity Near Point-of-Care Detection of Asymptomatic and Sub-microscopic Plasmodium Infections in African Endemic Countries"

[1] MRC Unit The Gambia at the London School of Hygiene and Tropical Medicine, Banjul, The Gambia
[2] Department of Infectious Disease, Faculty of Medicine, Imperial College London, London, UK
[3] ProtonDx, Translation & Innovation Hub, Imperial College London, London, UK
[4] Unité de Recherche Clinique de Nanoro, Institut de Recherche en Sciences de la Santé, Nanoro, Burkina Faso
[5] West African Centre for Cell Biology of Infectious Pathogens, College of Basic and Applied Sciences, University of Ghana, Legon, Accra, Ghana
[6] Faculty of Infectious and Tropical Diseases, London School of Hygiene & Tropical Medicine, London, UK
[7] Department of Electrical and Electronic Engineering, Imperial College London, London, UK
[8] Centre for Paediatrics and Child Health, Imperial College, London, UK
[&] These authors contributed equally

#### Table of Contents

**Supplementary Table 1**. Primer sequences of the LAMP assays used in this study ____ 2

**Supplementary Table 2.** Evaluating the dragonfly system specificity at various reaction time points ____ 3

**Supplementary Figure 1.** Confusion matrices for asymptomatic samples ____ 4

**Supplementary Table 3.** Comparison of the Dragonfly Pan/Pf malaria platform with malERA Target Product Profiles (TPP) Criteria for Community-Level Malaria Screening ____ 5

**Supplementary Table 4.** Comparison of the characteristics of the Dragonfly platform and the two commercial Malaria LAMP technologies ____ 7

**Supplementary Table 5**. Preliminary cost analysis of prototype approach, including both consumable and labour costs per sample, compared against market pricing for Alethia® Malaria Tests and required incubation and detection instrument ____ 8

**Supplementary Figure 2.** Giemsa-stained thin blood film of the final culture preparation, showing red blood cells (RBCs) singly infected with *Plasmodium falciparum* parasites at the ring stage ____ 9

**Supplementary Table 6**. Equations used to calculate sensitivity and specificity for values reported in the manuscript ____ 10

**Supplementary Methods** ____ 11

**Supplementary Table 1.** Primer sequences of the LAMP assays used in this study.

| Target | LAMP assay | Oligonucleotide sequence (5' to 3') |
|---|---|---|
| Pan/Pf | LAMP-PfK13_F3 | GGAGCAGCTTTTAATTACCTT |
| | LAMP-PfK13_B3 | ATGACATGAATTTAGAACTTCGCC |
| | LAMP-PfK13_LF | AATATGTTATGTTCATTATCAA |
| | LAMP-PfK13_LB | GAGAAAAAAATGAATTTTGGAGCT |
| | LAMP-PfK13_FIP | TGGTTGATATTGTTCAACGGAATCT-ATCAAATATATGTTGTTGGAGGT |
| | LAMP-PfK13_BIP | TGGCAATTTCTA AATGGTGTACCA-ATAAGAATCTGACAATGTGGC |
| | Polley-Pf_F3 | CTCCATGTCGTCTATCGC |
| | Polley-Pf_B3 | AACATTTTTTAGTCCCATGCTAA |
| | Polley-Pf_LF | CGGTGTGTACAAGGCAACAA |
| | Polley-Pf_LB | GTTGAGATGGAAACAGCCGG |
| | Polley-Pf_FIP | ACCCAGTATATTGATATTGCGTGAC-AGCCTTGCAATAAATAATATCTAGC |
| | Polley-Pf_BIP | AACTCCAGGCGTTAACCTGT-AATGATCTTTACGTTAAGGGC |
| | LAMP-Pan18s_F3 | GTATCAATCGAGTTTCTGACC |
| | LAMP-Pan18s_B3 | CTTGTCACTACCTCTCTTCT |
| | LAMP-Pan18s_LF | CGTCATAGCCATGTTAGGCC |
| | LAMP-Pan18s_LB | AGCTACCACATCTAAGGAAGGCAG |
| | LAMP-Pan18s_FIP | TCGAACTCTAATTCCCCGTTACC-TATCAGCTTTTGATGTTAGGGT |
| | LAMP-Pan18s_BIP | CGGAGAGGGAGCCTGAGAAA-TAGAATTGGGTAATTTACGCG |

**Supplementary Table 2.** Evaluating the Dragonfly system Specificity at Various Reaction Time Points.

| Reaction Time | True Negatives (TN) | False Positives (FP) | Total Negative Samples | Specificity (%) | 95% CI (%) |
|---|---|---|---|---|---|
| 50 min | 118 | 2 | 120 | 98.3 | 96.0 – 100 |
| 55 min | 118 | 2 | 120 | 98.3 | 96.0 – 100 |
| 60 min | 118 | 2 | 120 | 98.3 | 96.0 – 100 |

The analytical specificity of the Dragonfly Test was evaluated at different reaction time points. At 50 minutes, the assay demonstrated a specificity of 98.3% (95% CI: 96.0–100.6%), with 2 false positive results out of 120 negative samples tested. Extending the reaction time up to 60 minutes resulted in the same performance indicating the test maintains high specificity even with extended incubation periods over the maximum reading time.

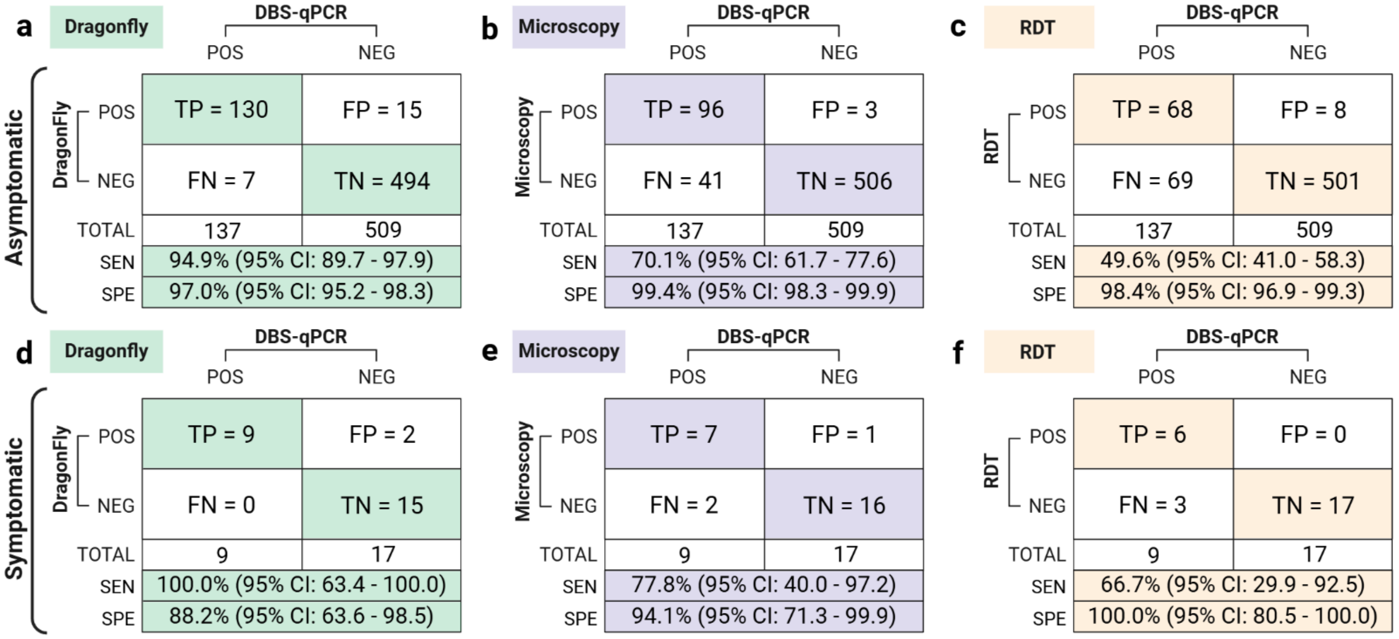


**Supplementary Figure 1.** Comparison of the clinical performance of Dragonfly, microscopy, and RDT methods using whole blood finger prick samples divided by asymptomatic and symptomatic cases, with DBS-qPCR as the gold-standard comparator. For each group, the number of true positives, total positive cases, sensitivity rate with 95% CI, number of true negatives, total negative cases, and specificity rate with 95% CI are provided.

**Supplementary Table 3.** Comparison of the Dragonfly Pan/Pf malaria platform with malERA Target Product Profiles (TPP) Criteria for Community-Level Malaria Screening

| **MalERA Criteria for Screening/Surveillance (District Level or Below)** | | **Status** |
|---|---|---|
| **Technical specifications** | | |
| Analytic sensitivity (parasite/µL) | E=20, D≤5 | |
| Diagnostic sensitivity | E>95%, D≥99% | |
| Analytic specificity | Negative all pathogens, common blood disorders | |
| Diagnostic specificity | E>99% surveillance low-transmission areas, E>95% screening | |
| Temperature stability | E, 30˚ C; D, 45˚ C for short periods | |
| Integrity of packaging | E, Moisture-proof | |
| **Species detection/differentiation** | | |
| P.f predominant areas | E, P.f; D, P.f/pan | |
| P.f and non-P.f areas | E, P.f/pan; D, differentiation all species | |
| Genotyping | No/O | |
| Ability to detect gametocytes | O | |
| Ability to detect hypnozoites | D | |
| **Health systems and technical specifications** | | |
| Packaging of tests or reagents | D, all required consumables enclosed; D, bulk packaging displays temperature violations | |
| Field stability/shelf life of consumables | E, 12 months (6 months since country); D, 2 y from manufacture (≥18 months in country) | |
| Training requirements | D, <1 week of pretrained medical technician | |
| Reagent requirements | E, nontoxic, all nonroutine provided; D, all necessary consumable items to perform the test provided in the kit | |
| Invasiveness | E, finger prick or less; D, non-invasive | |
| Rapidity of results | E≤2 days; D≤ half-day | |
| Ease of use | E, within medical tech ability; D, simple, few steps | |
| Cost | D≤US$1 per test | |
| Safety | E, high blood safety with basic universal precautions | |
| Waste disposal | Basic health system waste disposal | |
| Inter-reader reliability (clarity of result) | Kappa.0.9 | |
| Instrumentation and laboratory infrastructure requirements | D, all provided with test | |
| D = desirable, E = essential, O = optional | | |

| | | | |
|---|---|---|---|
| Desirable (D) criteria met | Essential (E) criteria met | Criteria not met | Data unavailable or not applicable |

**Supplementary Table 4.** Comparison of the characteristics of the Dragonfly platform and the two commercial Malaria LAMP technologies. (Loopamp™ Malaria and Alethia® Malaria)

| Characteristics | Alethia® Malaria | Loopamp™ Malaria | Dragonfly Malaria Pan/Pf platform |
|---|---|---|---|
| **General specifications and performances** | | | |
| Targets | Pan-Plasmodium | Pan-Plasmodium; *P.falciparum; P.vivax* | Pan-Plasmodium /*P.falciparum* |
| Analytical sensitivity for *Plasmodium falciparum* | 0.7-2 parasites/µL | 1–2 parasites/µL | 0.6 parasite/µL |
| Diagnostic sensitivity in symptomatic patients | 97.2 (92.6–99.1)[42] | 97.0 (89.6–99.6)[43] | 100 (63.4-100) |
| Diagnostic sensitivity in asymptomatic infected individuals | N/A | 83.3 (58.6-96.4) [a][40]<br>72.2 (62.6–80.2) [b][44]<br>40.8 (27.0–55.8) [c][41] | 94.9 (89.7-97.9) |
| Specificity | 87.7 (76.6–94.2)[42] | 99.7 (99–100) [a][40]<br>99.9 (99.8–100) [c][41]<br>99.2 (98.1–99.7 [43] | 96.8 (94.9-98.0) |
| Power requirements | Mains power | Mains power or portable battery | Mains power or portable battery |
| **DNA extraction** | | | |
| DNA extraction kit | Alethia Malaria Specimen Preparation or<br>Alethia Malaria PLUS Specimen Preparation | Loopamp™ PURE DNA Extraction Kit (optional)<br><br>Or boil &spin<br>Or other methods | SmartLid blood extraction Kit |
| DNA extraction technology | Filtration-based method | Heating step + adsorbent powder (Loo)pamp™ PURE DNA | Magnetic beads-based extraction |
| Sample type | Whole blood collected in EDTA | Whole blood or dried blood spots | Whole blood collected in EDTA |
| Sample volume | 50µl | 30µl | 100µl |
| Required equipment | None | HumaHeat Incubator [d] or HumaLoop M [e] | PDX Heat block or Regular Heat block |
| Weight of equipment | NA | HumaHeat :1.8kg | 1.03kg |
| Dimensions of equipment | NA | HumaHeat : 15cm × 17cm × 14.cm | 14cm*11cm*13cm |
| **DNA amplification** | | | |
| Required equipment | Alethia® Incubator Reader (Illumipro-10™ incubator/reader previously) | HumaLoop M [e] or HumaTurb A +C [f] | PDX Heat block or Regular Heat block |
| Weight of equipment | 2,95kg | HumaLoop M [e]: 9.5 kg or HumaTurb A+C [f]: 2.5 kg | 1.03kg |
| Dimensions of equipment | 21 cm x 29.2 cm x 9.5 cm | HumaLoop M [e]: 25 cm × 27.6 cm × 18.2 cm<br>HumaTurb A+C [f]: 52 x 35 x 23.5 cm | 14cm*11cm*13cm |
| Throughput<br>(maximum number of samples per run) | 10 | HumaLoop M: 16<br>HumaTurb A: 16 | 24 |
| Recommended incubation time | 40 minutes | 40 minutes | Min 20 minutes, Max 40 minutes |
| Reagents | Lyophilised | Lyophilised | Lyophilised |
| **Result readout** | | | |
| Detection method | Reaction solution absorbance characteristics | Fluorescence under UV light or turbidity | Colorimetric based on pH change |

| | | | |
|---|---|---|---|
| Required equipment | Alethia® Incubator Reader | HumaLoop M [e] or HumaTurb A+ C [f] | Naked eye |
| Output | Qualitative | Qualitative | Qualitative |

[a] Using boil & spin extraction method
[b] Using Chelex as extraction method
[c] Using HTP (High throughput) extraction method
[d] The HumaHeat Incubator is required when using HumaTurb A+C for amplification and result readout.
[e] The HumaLoop M integrates both sample preparation, amplification and result readout.
[f ] The HumaTurb A+C consists of two units: the HumaTurb A (amplification unit) and the HumaTurb C (control unit).

Comparison of Dragonfly with two Commercial LAMP Technologies. Dragonfly exhibits analytical sensitivity of 0.6 parasites/µL for *P. falciparum*, compared to 1-2 parasites/µL for Loopamp and 0.7-2 parasites/µL for Alethia. In symptomatic patients, Dragonfly demonstrated high diagnostic sensitivity (100%), comparable to Loopamp™ (97.0%) and Alethia® (97.2%)**.** For asymptomatic individuals, sensitivity data are only available for Dragonfly (94.9%) and Alethia (40.8-83.3%). All three platforms target either pan-*Plasmodium* or both pan-*Plasmodium* and *P. falciparum*, and can operate using mains power or portable batteries. Dragonfly requires only two regular heat blocks making it the lightest and most compact option. In contrast,  Loopamp and Alethia rely on dedicated incubators and readers, increasing their overall weight, size and cost. While market price per test is not yet available for Dragonfly, preliminary cost analysis estimates the prototype at £ 4.01 per test . Loopamp appears more cost-effective at approximately €5.2 per test, compared to Alethia at £ 43.

**Supplementary Table 5**. Preliminary cost analysis of prototype approach, including both consumable and labour costs per sample, compared against market pricing for Alethia® Malaria Tests and required incubation and detection instrument.

| **Component** | **Alethia® Malaria** *(approximate market cost, £)* | **Dragonfly Malaria Pan/Pf** *(estimated manufacturing cost, £)* | |
|---|---|---|---|
| Test cost per sample | 43[1] | Consumable test components (e.g. reagents, plastics, packaging, etc.) | 2.68 |
| | | Labour (UK-based, assuming batch sizes of <10K reactions) | 1.33 |
| | | Total test cost per sample | 4.01 |
| Instrumentation cost (e.g. for reaction incubation and result readout) | 16,000[2] | Isothermal heat block (for both sample lysis and LAMP incubation) | 112.00 |

[1] https://www.szabo-scandic.com/en/alethia-malaria

[2] https://www.fishersci.com/shop/products/alethia-incubator-reader/23029014

**Supplementary Figure 2**. Giemsa-stained thin blood film of the final culture preparation, showing singly infected red blood cells (RBCs) at the ring stage.

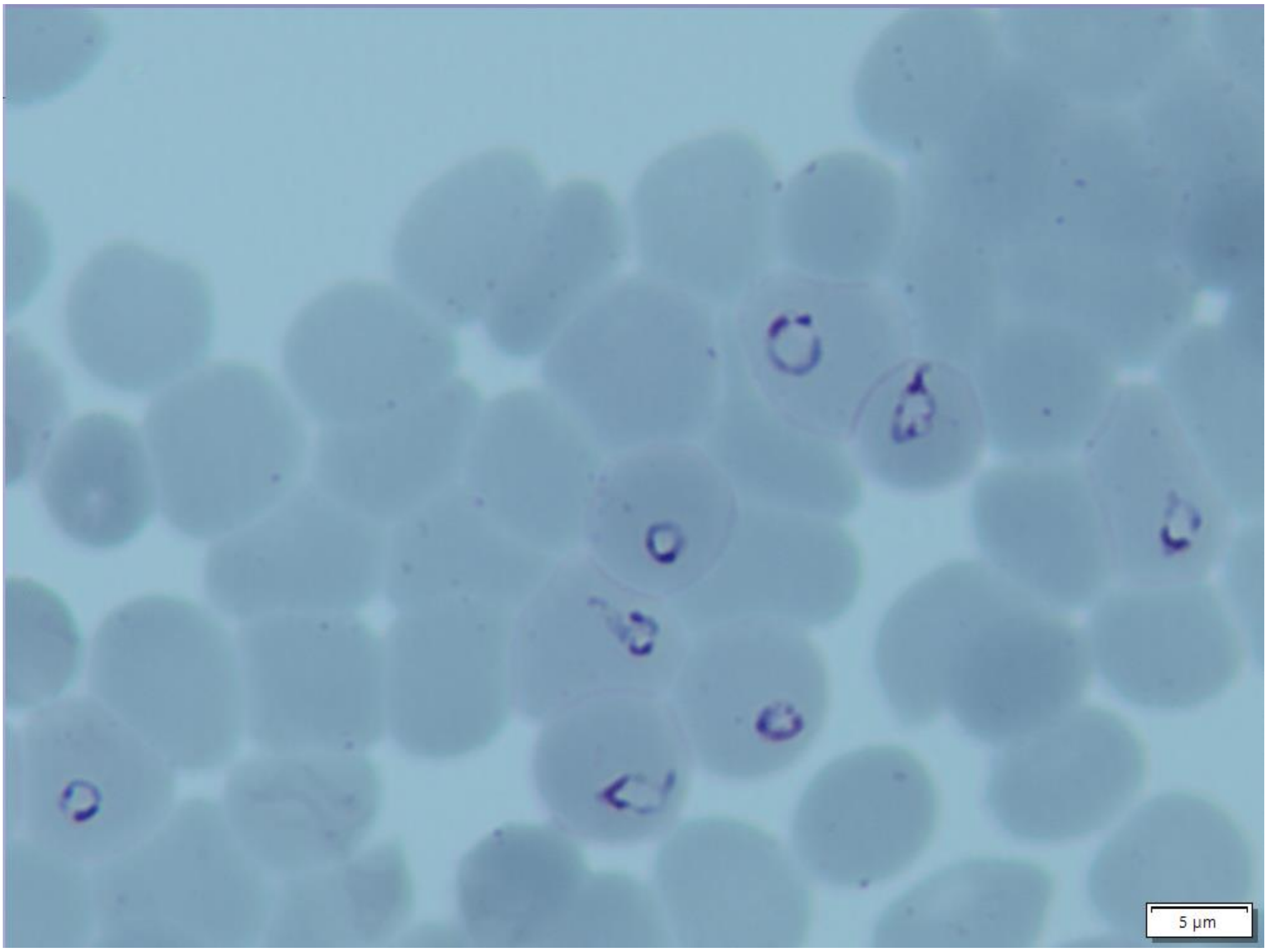

**Supplementary Table 6**. Equations used to calculate sensitivity and specificity for values reported in the manuscript.

| | **Comparator** | |
|---|---|---|
| **Test** | Positive | Negative |
| Positive | TP | FP |
| Negative | FN | TN |
| Total | $TP + FN$ | $FP + TN$ |
| Sensitivity | $\frac{TP}{TP + FN}$ | |
| Specificity | $\frac{TN}{TN + FP}$ | |

**Supplementary Methods**

### Protocol for high-throughput Extraction of DNA/RNA from Whole Blood with SmartLid and Detection with Dragonfly Malaria *Pan/Pf*

#### REQUIRED MATERIALS

- ProtonDx SmartLids
- ProtonDx Magnetic Keys
- ProtonDx 20 uL fixed volume Pipettes and tips
- ProtonDx Vortex mixer (optional)
- ProtonDx SmartLid Rack (optional)
- ProtonDx SmartLid Vortex Tool (optional)
- ProtonDx Heat block (locked to 63.5°C)
- Microcentrifuge tubes (2.0 mL)
- ProtonDx Lysis Binding Buffer
- ProtonDx Wash I concentrate
- ProtonDx Wash II concentrate
- ProtonDx Proteinase K
- ProtonDx Magnetic Beads
- ProtonDx Elution buffer
- Dragonfly Malaria pan/pf panels & result cards
- Molecular biology grade ethanol (>99%)
- Personal protective equipment PPE
- Decontamination solutions
- 1000 uL pipette and sterile filter tips
- Waste collection bag
- Timer
- EDTA collection tubes for whole blood

#### IMPORTANT Before Starting

- Always wear PPE, including lab coat and gloves. Change gloves if they touch anything not sterile.
- Prior to performing any biological procedure, ensure the working environment is clean using a decontamination spray according to local guidelines.
- Always use pipette tips that have a filter and are sterile.
- **Add 18 mL of molecular biology grade ethanol (>99%) to the Wash I (concentrate) bottle and 21 mL to the Wash II (concentrate) bottle** as indicated on the label.

#### SETUP for Multiple Extractions

1. Set up the required number of flip-cap tubes in a SmartLid Rack, with different rows for:

   **Lysis** (Row A)

   **Wash I** (Row B)

   **Wash II** (Row C)

   **Elution** (Row D)

   (For example, 6 extractions will require a total of 24 tubes, split in 4 rows of 6 tubes.)

2. If extracting from a **liquid sample**, including whole blood stored in EDTA tubes, prepare a master mix of Lysis Binding Buffer and Magnetic Beads in a 50 mL falcon tube according to the table below. Multiply the volume per tube by the number of extractions (n) and include an additional 10% to allow for pipetting errors:

| Component | Volume per tube [1] | Volume of Master Mix for n extractions |
|---|---|---|
| Lysis Binding Buffer | 600 µL | 600 µL × **n** |
| Magnetic Beads | 40 µL | 40 µL × **n** |
| **Total volume** | 640 µL | 640 µL × **n** |

[1] Use 10% overage calculation when making a master mix for use with multiple samples.

3. If needed, Negative extraction control (EDTA whole blood from healthy control) and Positive extraction control (EDTA whole blood spiked with malaria parasites) can be added for each user performing the Dragonfly test.

4. Prefill all tubes according to the table below:

| Tube/Step | Components and Volumes |
|---|---|
| A: Lysis Binding | **640 µL** Lysis Binding Master Mix |
| B: Wash I | **500 µL** Wash Buffer I [1] |
| C: Wash II | **500 µL** Wash Buffer II [1] |
| D: Elution | **50-200 µL** Elution Buffer |

[1] Ensure that molecular biology grade ethanol (>99%) has been added to the Wash Buffer (concentrate) as indicated on the bottle.

#### LYSIS BINDING

1. Add **20 µL** of Proteinase K to each Lysis tube (prefilled with Lysis Binding Buffer and Magnetic Beads master mix).

2. Add **100 µL** of whole blood to the Lysis tube.

3. Close with a SmartLid (without a Magnetic Key inserted) and briefly vortex the tube (**1-2 seconds**) to mix.

4. Incubate the lysis tube (with SmartLid inserted) at **65°C (or 63.5 if using ProtonDx Heat Block)** for **5 minutes**.

5. Remove the SmartLid and add **400 µL** of EtOH (>99%) to the lysed sample.

6. Return the SmartLid (still without the Magnetic Key) to the tube and pulse-vortex for **60 seconds**.

   **Note:** For processing multiple samples at a time, the SmartLid Vortex Tool (100174) can be used for each mixing step. Ensure the retainer disk is engaged securely before vortexing.

7. Insert a Magnetic Key into the SmartLid, turning it 90 degrees clockwise to lock, and invert the tubes several times to collect all Magnetic Beads onto the SmartLid.

   Allow the tube to remain upside down for **~30-60 seconds** after the first inversion, followed by multiple quicker (**~5-10 seconds each**) inversions to ensure all beads are collected.

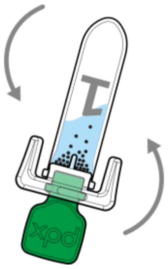

#### WASH

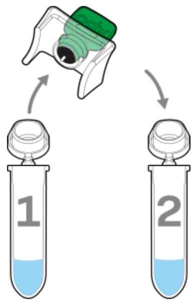


1. Transfer the SmartLid (with Magnetic Key INSERTED) from the lysis tube into the corresponding Wash I tube, ensuring it’s firmly inserted in the wash tube.

   **Important:** Ensure the Magnetic Key is inserted during transfer to avoid losing Magnetic Beads and captured nucleic acids.

2. Remove the Magnetic Key and pulse-vortex for **60 seconds** to resuspend the Magnetic Beads

3. Insert/lock the Magnetic Key into the SmartLid and invert the tube several times to collect all Magnetic Beads onto the SmartLid. **Ensure the liquid is clear before proceeding.**

   **Tip:** Initially shake the tube with the Magnetic Key inserted to first resuspend all Magnetic Beads, followed by gentle inversions for collection.

4. Repeat steps 1-3 for Wash II, transferring, resuspending, mixing, and again collecting the beads.

5. Once all washing steps are complete, and all Magnetic Beads are collected onto the SmartLid, remove the SmartLid and set it down (Magnetic Beads facing down) on a clean surface for **60 seconds** to **allow all EtOH to evaporate.**

#### ELUTION

1. Once the evaporation step is complete, insert the SmartLid (with Magnetic Key still INSERTED) into the corresponding Elution tube.

2. Remove the Magnetic Key and mix/shake the tube for **60 seconds** to fully resuspend the beads.

   **Important:** Do **NOT** use the vortex tool and vortex mixing for the elution step. Shake the tubes instead.

3. Insert/lock the Magnetic Key into the SmartLid. **Ensure the liquid is clear before proceeding.**

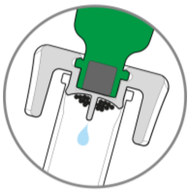

4. Flick the tube down to collect as much elution volume as possible, discard the SmartLid and attached Magnetic Beads, and store the elution tube. **Please do not discard the green Magnetic keys, as they can be cleaned and reused.**

#### TEST PANEL LOADING

Once the extractions are complete testing should be performed using the DF Malaria test panel (Pan/Pf) and dedicated heating block.

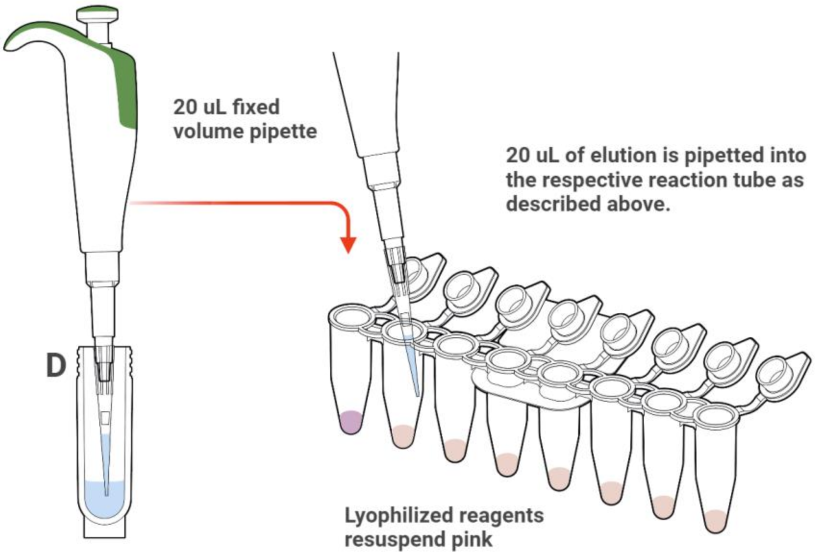

1. With a new pipette tip, rehydrate reaction **tubes 1 and 8** of the Pan/Pf test panels with 20 µL of the **Negative extraction control**, using the ProtonDx fixed volume pipette.  Discard tip.

2. With a new pipette tip, rehydrate reaction **tubes 2 to 7** with 20 µL of the elution's from **each patient sample**, making sure to **use a new tip for every sample**. Please consider each malaria strip can test up to 6 patients (P1 to P6) plus controls.

3. Flick down the Dragonfly reaction tubes to make sure all the liquid sits at the bottom.

   **Important:** Close lids securely!

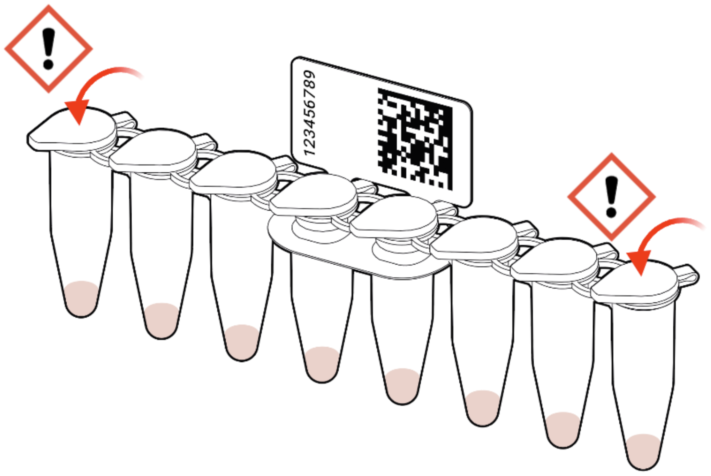


4. Transfer the DragonFly reaction tubes into the pre-heated ProtonDx heat block, after ensuring that the heater has reached 63.5°C making sure all the lids are securely closed.

5. Incubate the strip for 40 minutes.

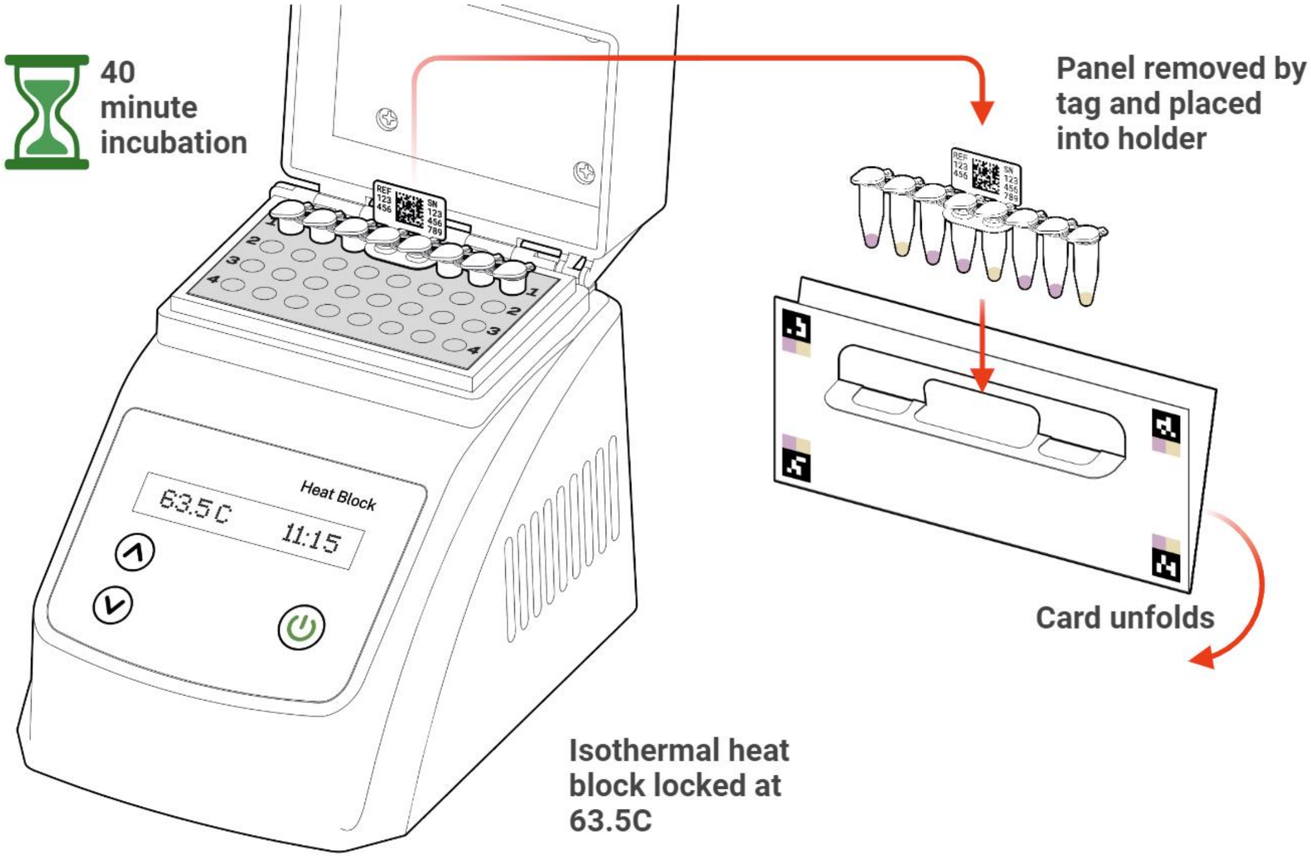

6. After the time has elapsed remove the DragonFly reaction tubes from the heater and place them on the results card.

7. Leave the strips to cool at room temperature for 10 seconds prior to taking a photo of the results.

#### RESULT INTERPRETATION

The user interpreting the panel should have good colour vision and be under good lighting conditions. A pen may be used to mark your selections and write additional details onto the card. **Write the Sample ID, Date, and operator's name on the card.** Insert the test panel into the card and fold open to show the result matrix.

**Important:** Always be sure the tube lids remain closed.

The Malaria Pan/Pf panel has 2 controls, and targets.

**Outcomes will be either Pink or Yellow.**

- The **tube 1** is a colour control and **must stay pink**. If it turns yellow, the test is invalid. Load the negative extraction control in tube 1.
- Tube 8 is an internal control and must turn yellow. The test is invalid if it remains pink.
- **Tubes 2, 3, 4, 5, 6, 7** are the targets and will be **either yellow or pink**. Yellow indicates the target is detected, or positive, and pink indicates the target is not detected, or negative.

**If the controls all match the expected colours, the test is valid.**

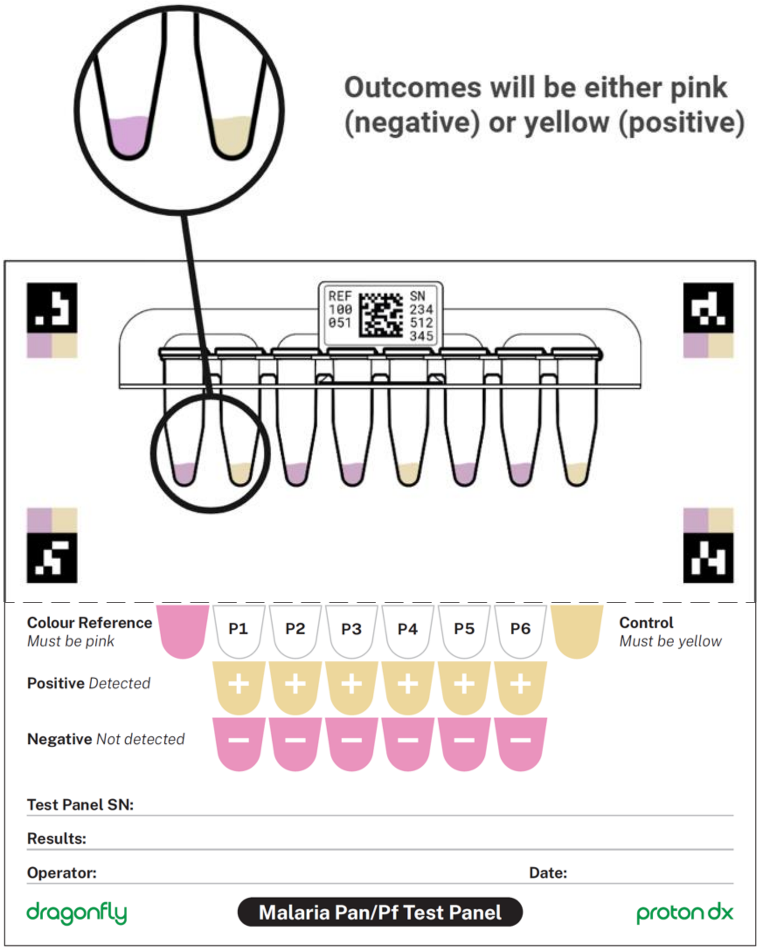
